# Beyond individual choice: a qualitative study on attitudes and social dynamics shaping pregnant women’s participation in clinical trials in Kano, Nigeria

**DOI:** 10.64898/2026.07.13.26357858

**Authors:** Mridula Shankar, Taiwo G. Amole, Faisal S. Dankishiya, Alya Hazfiarini, Fatima M. Mahmud, Muhammad G. Mukhtar, Abdulrahman I. Usman, Muhammad M. Bulama, Joshua P. Vogel, Sara Rushwan, A. Metin Gülmezoglu, Meghan A. Bohren, Hadiza S. Galadanci

## Abstract

**Background:** Nigeria’s high rates of maternal morbidity and mortality make it a crucial setting for maternal clinical trials. Little is known about how attitudes and social dynamics between communities and the health system shape pregnant women’s participation in such trials. This study explores these attitudes and social dynamics from the perspectives of women, their families, community leaders, health workers and policy-makers in Kano, Nigeria.

**Methods:** We conducted a qualitative study across 12 public health facilities at the primary, secondary and tertiary levels. We conducted 31 in-depth interviews with previous maternal health clinical trial participants, health workers and policy makers. We also held 11 focus group discussions with research naïve pregnant women, family and community members, and community health extension workers. Data were collaboratively analysed by the research team using reflexive thematic analysis.

**Results:** The participation of pregnant women in clinical trials was viewed by all participants as a strategy to mitigate pregnancy-related risks and adverse outcomes. Within social networks, women helped each other allay concerns about the likelihood and severity of participation-related harms through observation and discussion of positive experiences. Decision-making regarding participation was influenced by household gender dynamics, with husbands providing permission and financial support. Incentives for participation were valued, and when in monetary form, helped overcome some financial barriers such as transportation costs. Establishing and leveraging trust in health workers was key to successful recruitment and retention. Mechanisms for establishing trust were relational and included clear communication, respectful care and collaboration through a shared commitment to the participant’s health. As pivotal gatekeepers, health workers’ attitudes to clinical trial participation were shaped by their understanding of intervention efficacy and safety, and the potential impact of the trial on maternal health in Nigeria.

**Conclusions:** The participation of pregnant women in clinical trials in Kano, Nigeria, is shaped by intersecting social, cultural and economic factors. Collective decision-making, shaped by gender roles and social networks is critical to determining participation. Trust in health workers and positive experiences help alleviate concerns about participation-related harms. Strengthening local capacity and tailoring research processes to ensure cultural safety are essential for enhancing the conduct of maternal health trials.

## Background

The participation of pregnant women in clinical trials for medicines and medical product development is essential to identify new therapies and help reduce maternal and perinatal morbidity and mortality (1). At the same time, the conduct of clinical trials with pregnant women faces several unique challenges such as the altered pharmacokinetics of products due to physiological changes in pregnancy, additional ethical considerations and regulatory hurdles, and recruitment difficulties (2). Historically, concerns about fetal safety have led to the systematic exclusion of pregnant women from interventional research (3). This protectionist approach has created significant knowledge gaps regarding the safety and efficacy of medical product use in pregnancy to manage pre-existing conditions and conditions that arise in pregnancy (4). It has also obstructed the development of novel medications for preventing or treating obstetric conditions (5). These gaps have, in turn, contributed to suboptimal clinical care and avoidable risks and adverse outcomes for pregnant women and their babies (5).

Despite broad consensus that excluding pregnant women from clinical research causes harm, such exclusion is widespread (1). Factors operating at multiple levels of the research ecosystem contribute to the current situation. These include limited funder interest in novel obstetric therapies, ambiguities in regulatory and ethical review processes, liability concerns, sub-optimal healthcare infrastructure and human resources to support trial implementation, and complexities of trial design like determining appropriate endpoints for obstetric and neonatal outcomes (1, 6). Women may also have a low tolerance for risk during pregnancy, posing challenges to trial recruitment (6). However, synthesis of data on the views and experiences of pregnant women across high, middle and low-income countries suggests that pregnant women are willing to participate in clinical research when they perceive clear value such as health improvements for themselves or their babies, higher quality care, or contributing to greater societal good (6). Their willingness is strengthened when they trust the health system, receive support for participation from their families and communities, and experience trial procedures as acceptable and culturally safe within prevailing community norms, beliefs and practices (6). Understanding how community and health worker attitudes and socio-cultural influences shape decisions about trial participation in pregnancy in different geographical contexts is therefore critical to designing high-quality, ethically sound research that is feasible and acceptable.

The number of maternal and perinatal health trials conducted in low-and middle-income countries has steadily increased from 2010 onwards (7), alongside a growing emphasis on the inclusion of pregnant women in clinical research. Yet just 20% of these trials were in sub-Saharan Africa (7), despite the region accounting for approximately 70% of global maternal deaths (8). Nigeria, the most populous country in sub-Saharan Africa, is an important location for practice-changing obstetric trials (9–11). Nigeria’s maternal mortality ratio (an estimated 993 maternal deaths per 100,000 live births in 2023) has been stubbornly high with only a 14% reduction since 2000 (8). Nigeria accounts for more than one quarter of all maternal deaths globally, with Nigerian women facing a 1 in 25 lifetime risk of maternal mortality (8). This burden is disproportionately experienced by poorer women and households underserved by the health system, where structural inequities — such as limited access to quality antenatal, birthing and emergency obstetric care — greatly heighten their risk (12). Maternal deaths arise from both direct obstetric conditions such as haemorrhage, hypertensive disorders, and maternal infections, but also common conditions such as malaria, HIV, anaemia, and sickle cell disease that can endanger the lives of women during pregnancy (13–15).

Despite Nigeria’s growing prominence as a site for maternal and perinatal clinical trials, there has been no accompanying research exploring how women, their families, communities and health workers understand and experience clinical trial participation in pregnancy. In a context of high mortality, pronounced socio-economic and health disparities, and growing clinical trial activity, such evidence is vital to identify how social dynamics influence attitudes to trial participation. These insights can inform the design and implementation of future trials to ensure cultural congruence and ethical robustness. This study aimed to explore attitudes and social dynamics shaping the participation of pregnant women in clinical trials, drawing on the views of women, family members, community leaders and health workers in Nigeria.

## Methods

### Study design

This qualitative study was conducted within a constructivist paradigm to explore the attitudes, motivations and experiences of key stakeholders regarding pregnancy-focused clinical trials (16). Within a constructivist paradigm, it is understood that there can be multiple realities of a phenomenon, and interpretations of these realities are co-created through interactions between participants and researchers (17). Participant data were collected through focus group discussions and in-depth interviews. This study is reported according to the Consolidated Criteria for Reporting Qualitative Research (COREQ) (18) (Appendix 1).

### Study setting

This study took place in Kano, Nigeria from May 2024 to February 2025. We selected Kano because of its large population of reproductive-aged women, high burden of maternal mortality (1,147 maternal deaths per 100,000 live births (19)), and culturally embedded gender norms that impact women’s healthcare decisions (20). A key consideration in our choice of study setting was the role of the Amino Kano Teaching Hospital as an active maternal health research institution, prompting the inclusion of often underrepresented northern Nigerian populations in health research. We purposively sampled 12 public facilities representing the three-tiered structure of the Nigerian public health system and settings where clinical trials are conducted. Characteristics used to guide purposive selection of study sites (Table 1) included health facility level (primary, secondary, tertiary) and geographic location (urban, semi-urban and rural). We also purposively selected sites based on prior research exposure, including “research naïve” facilities with no previous clinical trial involvement, and “research experienced” facilities that had participated in at least one pregnancy-focused randomised controlled trial. We aimed to achieve representation across all strata rather than equal numbers in each, and the final distribution of sites reflects this aim and practical considerations such as feasibility and existing maternal health service provision patterns.

**Table 1.** Characteristics of sampled facilities (n=12)

| Characteristic | N (%) |
| --- | --- |
| **Clinical trial engagement** |  |
| Research-established | 7 (58.3%) |
| Research naive | 5 (41.7%) |
| **Level of health facility** |  |
| Primary | 3 (25.0%) |
| Secondary | 6 (50.0%) |
| Tertiary | 3 (25.0%) |
| **Type of health facility** |  |
| Public | 12 (100%) |
| **Geographic setting** |  |
| Urban | 8 (66.7%) |
| Semi-urban | 2 (16.7%) |
| Rural | 2 (16.7%) |
| **Average births per month (2023)** |  |
| <100 | 5 (41.7%) |
| 100–199 | 4 (33.3%) |
| 200-299 | 3 (25.0%) |

### Study participants, recruitment, and sampling

Participants were purposively sampled from the following groups: 1) pregnant women with no prior clinical trial experience (‘research naïve pregnant women’), 2) women who participated in a randomised controlled drug trial, either on anaemia in pregnancy (IVON trial) (11) or on testing a simplified magnesium sulphate regimen for eclampsia prevention (STEPMAG trial) (21) (‘research experienced women’), 3) health workers, 4) community health extension workers, 5) family and community members, 5) policymakers and, 6) ethics committee members. We used maximum variation sampling to access a range of experience levels and health worker types. Research findings from ethics committee members will be reported separately.

Recruitment occurred on-site at health facilities for research naïve pregnant women, family and community members, health workers, and community health extension workers. Clinical trial coordinators contacted previous trial participants to access research experienced women. Policymakers were approached directly by the research team, leveraging existing professional relationships. All participants received plain-language study information and provided written informed consent prior to participation. No potential participant declined participation.

### Data collection, study instruments, and data management

We conducted In-depth interviews with research-experienced women, health workers, and policymakers to explore motivations and experiences related to trial participation and implementation. Interviews lasted 30–60 minutes and were scheduled at the participants’ convenience. Focus group discussions were held with research-naïve pregnant women, mixed-gender groups of family and community members, and community health extension workers, to explore awareness and attitudes toward trial participation. Each focus group discussion included 6–10 participants, lasted 30–90 minutes, and was conducted in private, convenient locations within health facilities or communities. Two research team members were present at each in-depth interview and focus group discussion, serving as interviewer/facilitator and notetaker. In compensation for their time, research naïve pregnant women, family and community members, and community health extension workers received ⍰5,000 (USD$3.25) each; research experienced women and health workers each received ⍰10,000 (USD$6.50); and policymakers received ⍰20,000 (USD$13.00). We adapted and refined interview and discussion guides previously used in a related study in India (Shankar et al., 2025) (Appendix 2). Revisions included incorporating contextually appropriate terminology, such as a locally relevant definition of clinical trials with illustrative examples.

Data were collected in Hausa or English, audio-recorded, transcribed verbatim into English by members of the research team and anonymized by removing personal identifiers. Unique participant identification codes were assigned to each recording and transcript. Socio-demographic data were securely managed using KoBoToolbox (22). Electronic data were stored on password-protected computers, and paper records were secured in locked cabinets only accessible by research staff.

### Data analysis

The research team (MS, TA, FD, AH, FM, AI) conducted reflexive thematic analysis combining inductive and deductive approaches (23). An initial codebook, developed from a related study in India (24) was adapted for the Nigerian context. After data familiarisation through multiple readings, the first five transcripts were independently coded line-by-line by team members and discussed collectively over multiple team meetings to incorporate diverse analytical perspectives and positionalities. The codebook was refined, and remaining transcripts were coded individually, with ongoing team communication. These discussions were incorporated into the analysis and interpretation. Each team member updated the shared codebook as and when they developed new codes. Throughout this process, we communicated actively through a WhatsApp group and email to clarify and discuss the analysis.

We then held a two-day online workshop to discuss the grouping of codes into potential themes. We used Dedoose (25) to code, manage and review coded data, supporting collaborative team-based analysis.

### Researcher reflexivity

Our team of co-authors represents a diverse set of personal, geographical, social, and professional backgrounds, with members both with and without lived experience of pregnancy and involvement in perinatal clinical trial implementation. The core implementation team comprised 10 researchers: seven based in Kano, Nigeria; two in Melbourne, Australia; and one splitting time between Melbourne and Lombok, Indonesia. Collectively, our disciplinary expertise spans social sciences, epidemiology, medicine, and public health, with a range of experience applying qualitative methodologies in maternal health research. Many co-authors—including all those based in Kano—have also contributed to implementing obstetric trials as part of larger multi-country collaborations. This study was motivated by a shared commitment to exploring the factors shaping pregnant women’s participation in clinical trials in northern Nigeria, particularly through lenses of gender and health equity.

Throughout the project, we engaged in a reflexive process, considering and documenting how our disciplinary training, extent of foreignness and personal orientations influenced our engagement with the data. We also examined how our multiple intersecting social positions—including factors such as gender, age, geographic location, institutional affiliation, profession, and socioeconomic status—shaped our relationships to the data and the subsequent processes of coding and interpretation. These potential power and knowledge asymmetries were discussed and documented through a process of critical reflection.

For example, team members with clinical backgrounds recognised that research is inherently relational and reflected on the risk of being drawn more readily to the perspectives of health workers, which could inadvertently overshadow the views of women, their families, and community members. This awareness fostered open dialogue during the coding process to ensure balanced representation of all participant perspectives. Team members based at the University of Melbourne were also aware of the limits of their abilities to engage with the data in-context. These limitations were acknowledged and managed to the extent possible through data interpretation discussions led by team members in Kano.

## Results

Thirty-one In-depth interviews were conducted with research-experienced women (n=10), health workers (n=18), and policymakers (n=3). Eleven focus group discussions were held with research-naïve pregnant women (3 groups), family and community members (4 groups), and community health extension workers (4 groups). Characteristics of the participant groups are presented in Table 2. Most stakeholder groups were predominantly woman, except for family/community members. Younger pregnant women (20-29 years) were well represented, while older individuals (40+) were more prominent among influential roles such as family and community members, community health extension workers, health workers, and policymakers. Half of the research-experienced women had completed post-secondary education, while most research-naïve pregnant women graduated from primary and post-secondary schools. Most research-experienced women were multigravid, whereas most research-naïve pregnant women were primigravid. The median years of work experience were 9 years for policy makers and community health extension workers and 18.5 years for health workers.

**Table 2.** Sociodemographic characteristics of study participants (n=113)

| Characteristic | Previous research participants (n=10) | Research naïve pregnant women (n=23) | Family and community members (n=32) | Community health extension workers (n=27) | Health workers (n=18) | Policy makers (n=3) |
| --- | --- | --- | --- | --- | --- | --- |
| **Gender** |  |  |  |  |  |  |
| Woman | 10 (100.0) | 10 (100.0) | 9 (28.1) | 20 (74.1) | 13 (72.2) | 2 (66.7) |
| Man | 0 (0.0) | 0 (0.0) | 23 (71.9) | 7 (25.9) | 5 (27.8) | 1 (33.3) |
| **Age** |  |  |  |  |  |  |
| 15-19 years | 0 (0.0) | 1 (4.4) | 0 (0.0) | 0 (0.0) | 0 (0.0) | 0 (0.0) |
| 20-24 years | 2 (20.0) | 15 (65.2) | 1 (3.1) | 1 (3.7) | 0 (0.0) | 0 (0.0) |
| 25-29 years | 3 (30.0) | 1 (4.4) | 1 (3.1) | 6 (22.2) | 1 (5.6) | 0 (0.0) |
| 30-34 years | 3 (30.0) | 0 (0.0) | 4 (12.5) | 7 (25.9) | 4 (22.2) | 0 (0.0) |
| 35-39 years | 2 (20.0) | 5 (21.7) | 5 (15.6) | 1 (3.7) | 0 (0.0) | 1 (33.3) |
| 40+ years | 0 (0.0) | 1 (4.4) | 21 (65.6) | 12 (44.4) | 12 (66.7) | 2 (66.7) |
| Don't know | 0 (0.0) | 0 (0.0) | 0 (0.0) | 0 (0.0) | 1 (5.6) | 0 (0.0) |
| **Highest Educational level (completed)** |  |  |  |  |  |  |
| No formal schooling | 0 (0.0) | 4 (17.4) | 2 (6.3) | NA | NA | NA |
| Primary school | 0 (0.0) | 9 (39.1) | 2 (6.3) |  |  |  |
| Secondary school | 4 (40.0) | 1 (4.4) | 12 (37.5) |  |  |  |
| Post secondary school | 5 (50.0) | 9 (39.1) | 10 (31.5) |  |  |  |
| Islamiyya | 1 (10.0) | 0 (0.0) | 6 (18.8) |  |  |  |
| **Gravidity** |  |  |  |  |  |  |
| Primigravida | 1 (10.0) | 17 (73.9) | NA | NA | NA | NA |
| Multigravida | 9 (90.0) | 6 (26.1) |  |  |  |  |
| **Gestational age at time of data collection** |  |  |  |  |  |  |
| First trimester (1-3 months) | NA | 2 (8.7) | NA | NA | NA | NA |
| Second trimester (4-6 months) |  | 14 (60.9) |  |  |  |  |
| Third trimester (7-9 months) |  | 7 (30.4) |  |  |  |  |
| **Relationship to women participants** |  |  |  |  |  |  |
| Family member | NA | NA | 20 (62.5) | NA | NA | NA |
| Community member/leader |  |  | 12 (37.5) |  |  |  |
| **Health worker cadre** |  |  |  |  |  |  |
| Nursing staff | NA | NA | NA | NA | 9 (50.0) | NA |
| Medical officer |  |  |  |  | 5 (27.8) |  |
| Resident doctor |  |  |  |  | 2 (11.1) |  |
| Consultant doctor |  |  |  |  | 2 (11.1) |  |
| **Years of work experience: median (IQR)** | NA | NA | NA | 9 (2,27) | 18.5 (7,22) | 19 (6,20) |

Research findings are organised and presented in five themes: (1) research participation as a pathway to safer pregnancy, (2) participation decisions are mediated by male authority, (3) incentives support participant recruitment and retention, (4) trust is a precursor to recruitment and participation, and (5) health workers as catalysts for clinical trial participation.

### Research participation as a pathway to safer pregnancy

Within close-knit family and community structures in Kano, significant personal and social value was placed on pregnancy, childbirth and their outcomes. Shaped by this context, discussions about the value of trial participation were framed by the toll of maternal ill-health and death at a societal level, alongside the individual imperative to protect themselves from complications and possible fetal loss. For some research-naïve pregnant women, pregnancy was viewed as inherently risky — a situation in which a woman is “always sick”. Given this worldview, they welcomed trial participation as an opportunity to alter the course of potential ill-health and reduce the need for emergency interventions. As one participant explained:

> *“The reason why women will choose to participate in a clinical trial is, from the moment she conceives, she will start having some problems, like high blood pressure in pregnancy, anaemia…even eclampsia or prolonged labour which will lead to the woman having caesarean section. So, if she is told that there is a new drug that will alleviate all these things, she is likely to enter or participate in a clinical trial so as others may benefit from it.”* [focus group discussion, research naïve pregnant women]

This perspective was echoed by some family and community members who framed the individual and societal benefits of trial participation within the broader context of “a lot of suffering for pregnant women”. Previous research participants of the IVON and STEPMAG trials emphasised how their own experiences with potentially life-threatening conditions, as well as witnessing consequences among acquaintances, strongly motivated their participation. One woman, initially hesitant to enrol, described what led her to change her mind:

> *“At first, I even said I will not go, but because I had a problem during the time, I was in fear because there was a lot of mortality of pregnant women resulting from blood problems. So I decided to give it a try, because if you have anaemia while pregnant it is either of the two, whether to lose the baby or both the baby and the mother.”* [in-depth interview, previous research participant, IVON trial]

Concerns about the likelihood and severity of harms and side-effects associated with an experimental intervention were central to decision-making. As a previous research participant discussed:

> *“nothing is certain. It is a must to think that this may be harmful to you or your baby. It is only after participating that you will realise that, that is not the case.* [in-depth interview, previous research participant, IVON trial]

A community health extension worker noted that the novelty of an experimental drug can provoke “*fear and panic*” in pregnant women who bear responsibility not only for their own health but also for “*another life in you*.” These anxieties were shared by family members, whose involvement and responses influenced the final decision to participate or not. While some family members described offering encouragement and support by “*soothing her fears”*, other family members expressed active resistance to the very idea of participation, questioning “*are you insane? Why not allow others to enter* [the trial]”?

Imperfect knowledge and uncertainty about the intervention’s safety were neither static nor insurmountable. Where the risks of pregnancy and childbirth were visible within communities, social dynamics and learning through observation and discussion played a vital role in mobilising pregnant women to join trials. The experiences of women who joined trials first — driven by hopes of overcoming ill-health and protecting their babies — were closely watched and widely shared in social networks.

> *“If she participated in the clinical trial and she succeeded [her condition improved], this is the impact of the medication. So, with this success, she will tell other people (…) this is how the society will agree seeing one of them has participated and succeeded without any problem.* [focus group discussion, research naïve pregnant women]

When things went well, the process fostered trust and encouraged peers to enrol. As one community health extension worker explained, women were motivated towards a collective goal of reducing maternal mortality.

### Participation decisions are mediated by male authority

Gendered power relations, at multiple levels, shaped decision-making regarding trial participation. Several participants emphasised the gate-keeping roles of male authority figures in communities and households, who often mediated pregnant women’s consent and trial enrolment. As one community member put it:

> *“The people that can convince a woman to participate are, first her husband, secondly religious leaders, because if they say something and it goes viral, people really accept it.”* [focus group discussion, family and community members, research naive site]

At the community level, early and sustained engagement of male religious and community leaders through “*sensitisation*” was regarded as essential by all participant groups for a trial to gain acceptance. Their endorsement was seen as a key mechanism for building trust, and a prerequisite for the community to perceive the trial as credible and safe. As a medical officer explained:

> *“You have many religious leaders in the society; you have many district heads. Whatever they say is like authority. They are like demigods to people. So they [community members] follow because they have very strong impact and influence.”* [in-depth interview, health worker, research established site]

Faith leaders, leveraging their authority within religious institutions, could publicly engage men in places of worship. This reinforced support for a trial and shaped household-level decision-making. Participants described men’s control over such decisions about a woman’s trial participation as consistent with prevailing patterns of male authority over their spouses and daughters, and women’s deference to the male head of household.

> *“Sometimes after prayers they [community leaders] have a meeting with the husbands. If they convince the husbands that’s all [participation is acceptable], you know the women always follow their husbands. So when he tells her to go [participate in the trial], she will definitely go. The husband has the last words.”* [in-depth interview, health worker, research established site]

Husbands’ permission was frequently characterised as an expression of male guardianship over women. Trial participation often required travel and public interaction — doing so without a husband’s permission would challenge gender norms and risk adverse consequences. This might result in suspicion, marital conflict or, in extreme cases, threats of divorce. Gendered power relations over trial participation also intersected with structural factors, like poverty and education. Male control over household finances was the norm, hence a husband’s willingness and ability to provide resources — such as transportation fares — hinged on the household’s financial situation, and on his perception of whether trial participation was valuable. As a health worker noted:

> *“Some men will think they are just going for rubbish, or some men that they don’t trust their wives, they will think maybe they are just doing “yawo”* [unnecessary outing] *or doing all those “gulma”* [gossip]…*But if they are literate ones, so they know they will encourage them to… (and give them) financial support to even go there”* [in-depth interview, health worker, research naive site]

Some men struggled to meet their family’s basic needs and were less likely to support trial participation, especially if they had low literacy.

> *“Because some husbands are illiterate, they won’t even understand you. He will say this is the least of his problem, rather give him fertiliser if you really want to help him, or food to feed his family.”* [focus group discussion, research naïve pregnant women]

Poverty also influenced men’s concerns about their ability to manage potential harms associated with trial involvement. Some research-naive pregnant women noted that knowing their “*husband is financially stable*” helped ease these fears, while the absence of such financial security led to a decision not to participate.

Although less commonly discussed, both research-naive pregnant women and previous research participants highlighted the possibility — and occasional occurrence — of collaborative or autonomous decision-making. One previous research participant noted how she accepted participation “wholeheartedly” and signed a consent form before informing her husband, motivated by her trust in the recruiting nurse. Another woman described asserting that her own consent was sufficient, telling the nurse who was obtaining consent:

> *“my husband is at work, so even if I say yes now, he will not say otherwise because it is about my health”*. [in-depth interview, previous research participant, STEPMAG trial]

In focus group discussions, some research naive pregnant women expressed that joint decision-making was feasible, when wives could effectively communicate the details and benefits of participation to their husbands. This ability was sometimes described as being tied to structural factors, such as geographic residence and education. For example, educated women in urban areas were less likely to face restrictive gender dynamics at home, and tended to have greater involvement in decision-making.

### Incentives support participant recruitment and retention

The influential role of incentives for eligible participants was a recurring theme. Incentives for trial participants took various forms, including monetary payments, small gifts such as blankets or diapers, refreshments and drinks, or complimentary healthcare and medications. Regardless of type, incentives were consistently described as essential for reducing barriers to participation or enhancing the overall participant experience.

The poor state of the economy and household financial precarity affected the perceived value of monetary incentives. Such incentives were seen as crucial to overcoming some financial barriers to participation (particularly transportation costs) and also perceived as being valued by women for purchasing groceries or essentials for their children.

> *“Now with transport fare being very high, it is very easy for a woman to spend ⍰1,000 (approximately USD$0.65) in coming for clinical trial. Can you imagine her husband who is earning not more than ⍰2,000 or ⍰3,000 [per day], now to give her a whole ⍰1,000 for transport, it is not easy […] You can see the burden now, some of them they wanted to come but to come for follow-ups, unless there is support, which is why sometimes incentives for support or transport is so necessary that otherwise you are going to lose, some of them will get lost from the trial.”* [in-depth interview, health worker, research established site]

Material incentives, in the form of small goods, were also recommended. Some participants suggested that without some form of extrinsic motivation — material or monetary—women would be less likely to take part in a trial. Others identified free clinical care and medicines as incentives, particularly in situations where impoverished families would otherwise incur out-of-pocket costs. Incentives were frequently discussed among women within social networks, and served to raise awareness about the trial and encourage broader participation.

> *“Their enrolment in the trial really helps them, sometimes some of them cannot afford to take care of themselves, but because of the trial she wouldn’t have to worry about that and neither would her husband. They wouldn’t spend anything, since they are given incentives […] With that, a lot of women participate in clinical trials, because a woman tells another woman, like there is free medication at the hospital, there is even refreshments and incentives…”* [in-depth interview, health worker, research established site]

While incentives were generally viewed positively, some participants raised concerns about the optics of such incentives. In a focus group discussion with research naïve pregnant women, the use of incentives was met with suspicion and suggestive of veiled motives. This reaction was shaped by the lived experiences of women being denied care at health facilities, whereas when women are recruited for a trial, healthcare workers follow them home with incentives.

> *“Sometimes, even the incentive they will say there is some hidden agenda, because when you are sick and go to the hospital they will refuse you, but here the healthcare workers will be following you to your house with money.”* [focus group discussion, research naïve pregnant women]

Some health workers raised concerns about potential ethical issues related to undue inducement to participate. One health worker cautioned that revealing the incentive amount during informed consent could exert an undue influence on participants’ decision-making and potentially motivate others to participate for monetary reasons alone. Such details spread quickly through word of mouth. Another health worker spoke of the risk that trials might be perceived as purely profit-driven endeavours, with incentives seen in this light. To address this, they emphasised the need to explain to potential participants both the purpose of incentives and the potential health benefits of the trial.

> *“A lot of people now think a clinical trial is a means of getting money, so what will be their share. You have to inform them that this is not a business issue, there may be some incentives to help them with transport or something or to help them to have good rapport, but not that you are going to pay them something. Let them also understand that the information they are providing is going to assist many women who are coming subsequently, including herself if she continues to conceive.”* [in-depth interview, health worker, research established site]

### Trust is a precursor to recruitment and participation

> *“Because the level of trust between the health workers and the patient is so strong that whatever the health worker says, the patients obey.”* [in-depth interview, policy maker]

This quote from a policy maker conveys two linked concepts — trust and its relationship to power and vulnerability — which featured prominently in discussions about why women participate in trials. At an interpersonal level, building and maintaining trust in health workers was central to successful enrolment and retention. Trust was built (signified by bolded text below) through relational mechanisms and closely linked to how vulnerability and power imbalances between participants and health workers were navigated.

Participants described the importance of verbal and non-verbal **communication** across multiple stages of interaction. At enrolment, this meant taking the time to explain a trial’s purpose, procedures, risks and benefits. A friendly demeanour, clear and accessible language, respect and transparency were seen as vital for fostering trust. Trust was strengthened through **respectful care** when participants were offered comprehensive information in a compassionate manner. A previous research participant explained:

> *“Well, what I like about it is that the people in charge of it are kind-hearted and they tolerate somebody (…) No matter how you are [how you behave], no matter what you do, they will still try as much as possible to calm you down, and make you understand why this is happening and why that is not happening, I like their tolerance seriously.”* [in-depth interview, previous research participant, STEPMAG trial]

Regard for a person’s dignity, even when not fully compliant with trial procedures was particularly noted as fostering greater trust. As a previous research participant explained *“There was no problem even if I was late, no shouting or mistreatment. They understand if you have an excuse*.”[ in-depth interview, previous research participant, STEPMAG trial]

Health workers also built trust by developing **collaborative relationships** with participants and demonstrating shared investment in their health. These relationships were often established independent of trial implementation during routine health service delivery. Yet they were critical to allay concerns regarding the potential for harm. As a community health extension worker explained:

> *“We are the backbone of the whole thing, because we are the link between the researchers and the participants. So, if you have good contact with them and can counsel very well, that will definitely help (…) It is very important, because the women know and trust us, so they assume because we help solve their problems, we won’t bring something that will harm them.”* [focus group discussion, community health extension worker, research established site]

Some participants described trust between health workers and women in paternalistic terms. They described how deference to authority and an obligation to comply with medical advice shaped trial participation. In these accounts, relational dynamics tended to reinforce, rather than reduce, existing power and knowledge imbalances. As one community health extension worker explained:

> *“As people with some knowledge about these things [clinical trials], because whenever people come to the hospital and see you in uniform, they will regard you and accept whatever you tell them. Because they came with problems and you have them solved. So, this relationship will enable them to participate in a clinical trial.”* [focus group discussion, community health extension worker, research established site]

Some previous research participants also spoke of broader trust in the health system and their belief that these institutions protected their health. With this understanding, trial participation was described as an act of cooperation, arising from trust in the system to provide appropriate and safe treatment for ill-health. Conversely, discussions of mistrust in the health system were related to fears of harmful and extractive colonial practices. As one nurse explained:

> *“Some of them (…) would say the western people will never give you anything for free, there must be something in for them. That it may kill you or make you barren”* [in-depth interview, health worker, research established site]

When asked why some pregnant women might refuse participation, community health extension workers spoke of the spread of “*rumour*s”, especially influenced by family and community members. For example, they described people refusing immunisation programmes, such as Polio or COVID-19, due to fears that these will “*reduce your lifetime.*”

Health workers also described mistrust by explaining how community members make sense of family planning initiatives. Some people are concerned that these programs “*may be just like a pro-white people or pro-European program, that they want to cut the population of the state or of the country, just like a belief.”* [in-depth interview, health worker, research naïve site]. One community health extension worker noted that a clinical trial may be viewed as “*family planning in disguise.*” While expressed by a minority of participants, these accounts reveal that, for some, public health programs, are perceived as tools of neocolonial conquest.

### Health workers as catalysts for clinical trial participation

Health workers were seen as pivotal gatekeepers, with significant influence over patient decision-making and opportunities to join a trial. Health workers’ engagement was closely linked to their own knowledge and personal attitudes to pregnancy research. These attitudes were, in turn, shaped by the perceived potential of the trial to address obstetric challenges. As a resident doctor explained:

> *“There are so many factors now, because most of the trials are done somewhere, maybe outside even our continent, not even our country. So (…) I would like to see the effect in my population actually, because we have different demographics, different genetic composition.”* [in-depth interview, health worker, research experienced site]

Perceptions of safety were informed by the type of intervention being trialled. While some doctors acknowledged the underrepresentation of pregnant women in clinical research, they were generally sceptical of trials of novel medications or drugs not otherwise used in pregnancy. In contrast, they positively viewed interventions of treatment or care bundles, or trialling different doses of medications already in use during pregnancy.

Separately, some doctors raised ethical concerns about enrolling pregnant women in placebo-controlled trials, when, they considered the investigational drug as already safe in pregnancy. They felt discomfort with the idea that their “*patient may end up having the placebo*”, denying them access to a treatment needed “*as part of her management*”. A senior resident doctor expressed a separate ethical concern related to the dual role of clinician-researchers. In his view, when patients were unable to clearly distinguish between the roles of caregiver and researcher, there was a risk that they may “*think that if she does not accept what you offer her, in terms of involvement in a trial, she may think that you may not continue taking care of her the way you should.”* He was concerned these blurred roles could undermine voluntary participation or compromise ethical standards.

Health workers were keen to receive ongoing training on trial implementation. They viewed clinical research as critical to improving the health and survival of pregnant women and their babies. However, one doctor emphasised that meaningful engagement in research should also foster local leadership and build capacity to design contextually relevant trials. As he explained:

> *“Most of the trials that I was involved in are not locally designed (…) All of them are designed outside the country so it is when after they finish designing the project and everything, they will sit down and look at how are we going to customise whatever they design so that it will suit our environment and our place of work. So I would like to have more refresher courses on designing trials and executing them.”* [in-depth interview, health worker, research experienced site]

## Discussion

This study explores factors affecting participation of pregnant women in clinical trials, at the intersection of communities and the health system in Kano, Nigeria. We found that clinical trial participation was viewed as a means to address pregnancy-related risks and harms at both individual and societal levels, with collective decision-making shaped by strong gender roles and norms. Incentives for participation served as extrinsic motivators, by helping overcome financial barriers to research participation. Concerns about uncertainty and severity of harms were eased by observing trial experiences among early participants within social networks, and by fostering trust in health workers through respectful care. Health workers’ support for clinical trials in pregnancy was influenced by their belief in the potential for the intervention to address obstetric complications, and their confidence in intervention efficacy and safety within their local contexts. These findings align with factors demonstrated to operate at the interface between health systems and communities and extend current evidence by demonstrating how decisions regarding participation are embedded in kinship structures and shaped by experiences and influence within women’s social networks.

The research findings suggest that value ascribed to clinical trial participation during pregnancy must be considered within the broader context of persistently high maternal and infant mortality, where consequences of loss are felt deeply across communities. Our study shows how male gatekeeping roles, physical proximity of extended families, strong intergenerational bonds, and high social importance placed on pregnancy and childbirth converge within a collectivist culture to influence attitudes to and decisions regarding trial participation. These factors shape and interact with structural determinants of health such as poverty, gender inequality, and a history of colonialism to influence understandings of and responses to opportunities for clinical trial participation.

Informed consent is designed to embody the principle of respect for autonomy in decision-making (26). In practice, the educational and communicative process leading up to signing an informed consent form is shaped by a complex interplay of social relationships, gendered power dynamics, and broader structural factors, such as poverty. These interacting factors influence how pregnant women negotiate agency, power (or lack thereof), and decision-making around their participation. We recommend conducting formative research studies in diverse settings to improve understanding of how informed consent procedures for clinical trials should be adapted to optimise the relevance and suitability of the underlying ethical constructs to local cultural and contextual conditions. Previous research exploring this question underscores the importance of flexibility and openness to adaptations in the consent process to promote cultural safety and congruence (27, 28). Our findings reinforce these recommendations and offer insight into how male gatekeeping roles may require carefully negotiated, context specific strategies. However, it is essential that such adaptations do not undermine or override autonomy—for example, by substituting a woman’s own consent with that of her husband or family member. Designing research procedures to be culturally congruent and ethically sound require dedicated time and effort to building genuine cooperation and cultural understanding by working closely in the formative phase with local investigators, research staff, and ethics committees in study locations. This is particularly vital in multi-country trials that are led, sponsored, or bound by the institutional requirements of investigators or ethics committees in high-income countries, where alignment of standardised procedures with local circumstances may otherwise be overlooked.

Structural factors - such as poverty and its intersection with gender norms that restrict mobility and reinforce male control over household finances – produce inequities that constrain the ability of pregnant women to participate in research, and thus influence which groups are (and are not) represented in clinical trials. For instance, if the benefits of trial participation are perceived by the household decision-maker as uncertain, or less pressing than daily necessities, participation is deprioritised. Conversely, if a socially marginalised community sees trial participation as an opportunity for a pregnant family member to access healthcare that would otherwise be unattainable, this group may be overrepresented. These dynamics have implications for the ethical principle of justice, regarding the fair distribution of research risks and benefits. Some financial barriers to participation can be addressed through offering monetary incentives (29, 30) to cover the participants’ travel costs or lost wages.

Consistent with previous research, our findings highlight the significant influence of interpersonal trust in health workers, fostered and sustained through respectful caregiving (6, 24, 31, 32). However, leveraging health worker-patient relationships to encourage participation has also been described as a form of explicit or implicit compliance with health worker authority, reflecting a dynamic of dependency or involuntary trust (33). This dynamic can reinforce existing knowledge and power asymmetries in ways that are detrimental to promoting voluntariness. Transforming entrenched power hierarchies can be challenging. Nevertheless, pregnant women in this study supported approaches that strengthened relational trust—such as more compassionate, less prescriptive communication, and respectful care. Demonstrating a shared commitment to participant well-being, regardless of whether women choose to enrol, was also highly valued.

A particular challenge in trial research is the blurred distinction between research and clinical care, especially when health workers providing clinical care are involved in research implementation. The trust that develops within clinical care settings may be used, intentionally or not, to encourage trial enrolment. Potential participants may not feel comfortable declining participation or not fully comprehend the difference between clinical care and research, especially when participation offers access to free or higher-quality clinical care. In settings with high perinatal mortality, both health workers and participants may view trial participation as a valuable opportunity to reduce risks of ill-health or the severity of maternal, fetal, or newborn illness. This perception, combined with the trust built in caregiving relationships, can strongly influence how enrolment is valued.

Some participants in this study spoke of community mistrust towards the health system, shaped by fears that their lives may be regarded as expendable to serve the interests of those with racial privilege. These realities call attention to the need to critically examine how historical and current day racial, social, economic and political injustices influence ongoing relationships between communities and institutions in post-colonial countries (34). Clinical trials and public health initiatives —often indistinguishable to community members — are predominantly conceptualised, funded and directed through institutions based in high-income countries, reflecting donor priorities, and carrying legacies of extractive colonial practices (35). Acknowledging community mistrust as a protective response to invitations for trial participation is vital alongside addressing the need for fit-for-purpose therapeutics in pregnancy in settings with the highest burden of perinatal ill-health and mortality. Even within the constraints imposed by broader socioeconomic and political contexts, there are opportunities to take intermediate actions that reduce power asymmetries and shift power towards greater local ownership. Health workers in this study emphasised the unique value of locally designed trials in addressing pressing perinatal health concerns in their contexts. They expressed a strong interest in strengthening their competencies to lead trials from the ground-up. These findings complement calls for decolonising global health research and provide concrete entry points — such as trial training hubs and country-led protocols – for shifting power towards local researchers. Initiatives such as the ClinOps program is one model of promoting country ownership of clinical trials in sub-Saharan Africa (36). Through a structured training program, ClinOps aims to develop clinical trial training hubs that build local capacity and foster sustainable expertise within the region.

Our study has both limitations and strengths. In the focus group discussions with research naïve pregnant women and family and community members, some participants initially interpreted questions within the context of clinical care in pregnancy, rather than clinical trials. However, this challenge was anticipated, and we addressed it by developing a standard definition of clinical trials and using examples to regularly remind participants about the distinction. Our data did not explicitly explore participants’ understanding and views on specific trial design features, such as randomisation, which merits future investigation, particularly in relation to expectations of care. A key strength lies in engaging a diverse range of stakeholders at the intersection of community and health systems. This approach enabled data triangulation by highlighting where data converged or diverged within and across our participant groups. Including both research-naive and research-experienced participants allowed exploration of perspectives on and experiences participating in clinical trials. Our coding approach provided individual team members with autonomy to develop codes within an agreed framework, while regular interdisciplinary team discussions enabled critical reflection and multiple insights grounded in both disciplinary expertise and cultural understanding of context. Together, these elements contributed to the rigor of our analyses.

This study focused on exploring community and health worker perspectives on clinical trial participation in pregnancy, reflecting the specific physiological, ethical and practical considerations that shape trial design and participation during this period. Lactating women are also under-represented in drug and vaccine clinical trials, leading to gaps in evidence for the prevention and treatment of conditions during breastfeeding. Future qualitative research to understand how communities and health workers perceive clinical trial participation during lactation would help inform the responsible inclusion of lactating women in clinical trial research.

## Conclusions

We identified an interplay of social, cultural and economic factors influencing pregnant women’s participation in clinical trials in Kano, Nigeria. Collective decision-making, shaped by strong gender-based decisional dynamics, were central to research engagement. Incentives, experiences of participation in social networks, and trust in health workers helped address barriers and concerns about risk. Our findings emphasise the need for formative research to inform clinical trial designs that are responsive to local socio-cultural and health system contexts and to structural inequities, including tailored consent processes and recruitment strategies that recognise household and community roles in decision-making while still supporting women’s relational autonomy. They also highlight the importance of culturally congruent, sustained community engagement to build trust in contexts marked by collective concern about high maternal and infant ill-health, stark socio-economic disparities, and historical mistrust of externally driven research. Strengthening local research capacity and attention to improving cultural safety will be key to expanding maternal health-focused trials in this setting.

## Data Availability

The datasets generated and/or analysed during the current study are not publicly available to protect the identity of the participants but are available from the corresponding author on reasonable request. Please direct all data enquiries to the corresponding author.

## Declarations

## Ethics approval and consent to participate

This study received ethics approval from: The Kano Ministry of Health (NHREC/17/03/2018), the Research Ethics Committee, Amino Kano Teaching Hospital (AKTH/MAC/SUB/12A//P-3/VI/3859), and the Human Research Ethics Committee, University of Melbourne (2024-26117-52866-6).

## Consent for publication

All participants in this qualitative study consented to the publication of findings.

## Competing interests

None

## Funding

The research in this publication was supported by funding from MSD, through its MSD for Mothers initiative and is the sole responsibility of the authors. MSD for Mothers is an initiative of Merck & Co., Inc., Rahway, NJ, U.S.A. MAB’s time is supported by an Australian National Health and Medical Research Council Investigator Grant (2025634) and a Dame Kate Campbell Fellowship. The founders were not involved in the research process or interpretation of results.

## Authors’ contributions

AMG and MAB acquired funding for this study. All authors conceptualised and designed the study. Data collection was supervised by MG, MM and FD. MS, TA, FD, AH, FM, AI and MG conducted analyses. MS prepared the original draft and subsequent revisions with input from MAB, TA, FD, AH, FM, AI and MG. MAB and HG provided overall supervision for the project. All authors critically reviewed the manuscript including all revisions and agreed on the final version for submission to the journal.

## Acknowledgements

We acknowledge all the research participants who generously made this work possible. We are grateful to the project staff who collected and transcribed data for this study. We thank the Kano State Ministry of Health and Kano State Primary Healthcare Management Board for use of the health facilities to conduct data collection activities.

## Notes

### Competing Interest Statement

The authors have declared no competing interest.

